# COVID-19 Demographics, Acute Care Resource Use and Mortality by Age and Sex in Ontario, Canada: Population-based Retrospective Cohort Analysis

**DOI:** 10.1101/2020.11.04.20225474

**Authors:** Stephen Mac, Kali Barrett, Yasin A. Khan, David MJ Naimark, Laura Rosella, Raphael Ximenes, Beate Sander

## Abstract

**Background:** Understanding resource use for COVID-19 is critical. We conducted a population-based cohort study using public health data to describe COVID-19 associated age- and sex-specific acute care use, length of stay (LOS), and mortality.

**Methods:** We used Ontario’s Case and Contact Management (CCM) Plus database of individuals who tested positive for COVID-19 in Ontario from March 1 to September 30, 2020 to determine age- and sex-specific hospitalizations, intensive care unit (ICU) admissions, invasive mechanical ventilation (IMV) use, LOS, and mortality. We stratified analyses by month of infection to study temporal trends and conducted subgroup analyses by long-term care residency.

**Results:** During the observation period, 56,476 COVID-19 cases were reported (72% < 60 years, 52% female). The proportion of cases shifted from older populations (> 60 years) to younger populations (10-39 years) over time. Overall, 10% of individuals were hospitalized, of those 22% were admitted to ICU, and 60% of those used IMV. Mean LOS for individuals in the ward, ICU without IMV, and ICU with IMV was 12.8, 8.5, 20.5 days, respectively. Mortality for individuals receiving care in the ward, ICU without IMV, and ICU with IMV was 24%, 30%, and 45%, respectively. All outcomes varied by age and decreased over time, overall and within age groups.

**Interpretation:** This descriptive study shows acute care use and mortality varying by age, and decreasing between March and September in Ontario. Improvements in clinical practice and changing risk distributions among those infected may contribute to fewer severe outcomes among those infected with COVID-19.

## Introduction

Understanding the local, context specific, epidemiology and resource use implications of COVID-19 is critical to inform mitigation strategies for the second wave and throughout the pandemic. Allocating acute care resources appropriately and adequately for all patients and the ability to use targeted public health measures to minimize adverse effects resulting from broad restrictions is a key concern.(1–3)

Ontario population-level studies to date describe several aspects of the first wave: age and sex-specific descriptive studies for testing, cases and outcomes up to May 26, 2020,(4) and for hospitalizations up to June 17, 2020,(5) mortality using cremation data in a time series up to June 30, 2020,(6) and prediction tools using cases up until May 15, 2020.(7) However, as the COVID-19 pandemic evolves, current data on health outcomes and acute care resource utilization across stages of the pandemic are warranted.

The objective of our study was to describe COVID-19 cases in Ontario between March 1, 2020 and September 30, 2020, and to provide estimates of age- and sex-specific acute care resource utilization (hospitalization, ICU admission, invasive mechanical ventilation (IMV)), length of stay (LOS), and mortality.

## Methods

### Study Design, Setting and Participants

We conducted a population-based cohort study using administrative data collected from Ontario’s case and contact management (CCM Plus) database. We obtained research ethics board approval from the University of Toronto. CCM Plus is Ontario’s province-wide population-based dataset on all individuals who test positive for COVID-19 in Ontario (74,715 individuals between January 23 and October 30, 2020). (8) CCM Plus includes individual-level data on demographics (e.g., age, sex, region), epidemiology (e.g., likely acquisition), patient characteristics (e.g., co-morbidities), acute care resource utilization (e.g., hospitalization, ICU admission, IMV), health outcomes (e.g., mortality), and long-term care (LTC) residency. Given the evolving nature of the dataset, including addition of variables over time, our analysis was limited to outcomes of interest using fields which were considered complete by the dataset custodian. All variables used for this analysis are described in Appendix 1.

We accrued incident cases of laboratory-confirmed COVID-19 cases between March 1 and September 30, 2020 and followed these individuals until October 30, 2020, ensuring at least 30 days of follow-up. Accrual was based on the “accurate episode date” (episode date) field in CCM Plus. Since 94% of all cases require up to 14 days for the episode date to be completed (Appendix 2), and time from episode date to hospitalization is approximately 8 days, we only accrued cases until September 30, 2020 to allow for at least 30 days of follow-up.

### Outcomes

We examined acute care resource use (hospitalization, ICU admission, IMV), LOS at each level of acute care, and mortality. Outcomes are examined overall, by age and sex, specific co-morbidities, LTC residence status, and by month based on episode date. We considered three co-morbidities: diabetes, immunocompromised, and renal conditions, which were previously identified as conditions that increase risk of mortality among COVID-19 patients. (7)

### Analysis

We describe overall acute care use by 10-year age groups, sex, and month based on accurate episode date, which uses a number of dates entered to provide an approximation of onset date. Outcomes are calculated as follows: hospitalizations based on the total number of infections, ICU admissions based on the total number of individuals hospitalized, and IMV based on the total number of ICU admissions. We assumed that individuals who were recorded as “intubated” received IMV.

Mortality and LOS were estimated by acute care level: 1) Ward (i.e., hospitalized but did not receive ICU care or IMV), 2) ICU (i.e. required ICU care but no IMV) and 3) Ventilation (i.e., required IMV). Mortality was also estimated for individuals who were never hospitalized. For these outcomes, we only included individuals with resolved outcomes (resolved or fatal). We assumed death was attributable to the highest level of care (i.e., related to the severity of the disease). For example, if an individual was hospitalized, we do not differentiate whether this individual died during or after their hospitalization. Overall mortality was analyzed by month based on episode date.

Individuals missing data due to lack of follow-up or inaccurate dates (e.g., hospitalization admit dates entered as 2018, 2019, early 2020) were excluded from the analysis. Detailed information on data manipulation and cleaning are described in Appendices 3-5.

All data was handled and analyzed in Microsoft Excel 2016. Results are reported following the RECORD statement for observational studies (Appendix 6). (9)

## Results

### Demographics

Until October 30, 2020, there were 75,715 individuals with laboratory confirmed COVID-19, of which 10 were excluded due to incomplete data, and 18,215 did not meet the accrual period (56 pre-accrual, and 18,169 post-accrual), resulting in a total of 56,476 individuals from March 1 to September 30, 2020 included in this analysis. Approximately 64% of cases were between 20 and 59 years of age, and 52% were female. Approximately 18% of all cases were 70 years old and older. When cases associated with LTC residency are excluded, the proportion of cases among those age 70 years and over drops to 9%. Among LTC residents, 87% were 70 years or older, and 64% were female. Characteristics of all cases are summarized in Table 1.

**Table 1.**
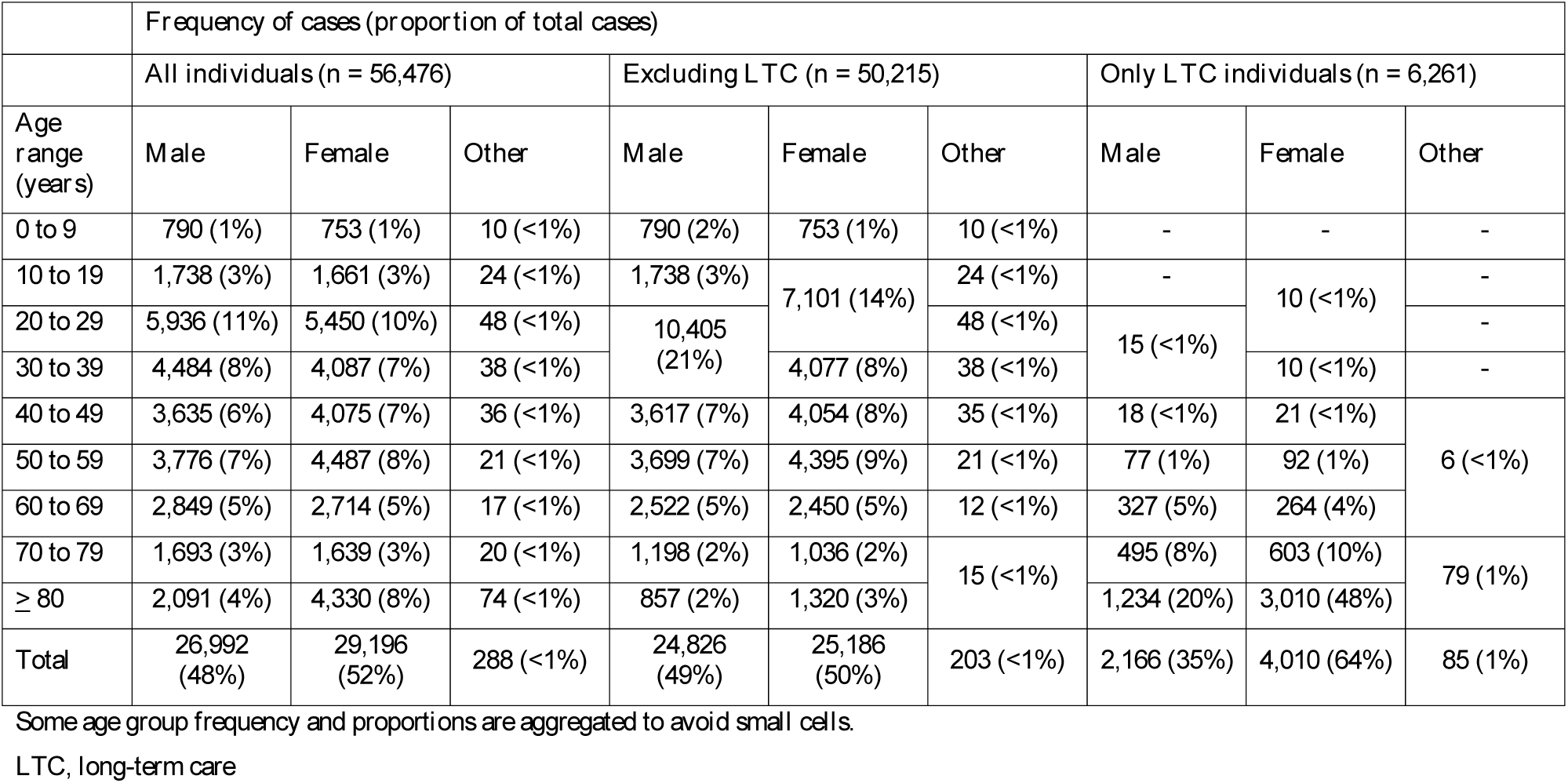
Demographics for individuals infected with COVID-19 between March 1 and September 30, 2020.

Figure 1 shows total cases by age group over time overall (Figure 1A) and excluding LTC residents (Figure 1B). As the pandemic progressed, the proportion of total cases from older population groups (≥ 60 years) decreased from a high of 46% in April to 13% in September, while the proportion of total cases from younger populations (age 10-39 years) increased from a low 25% in April to 55% in September.

**Figure 1.**
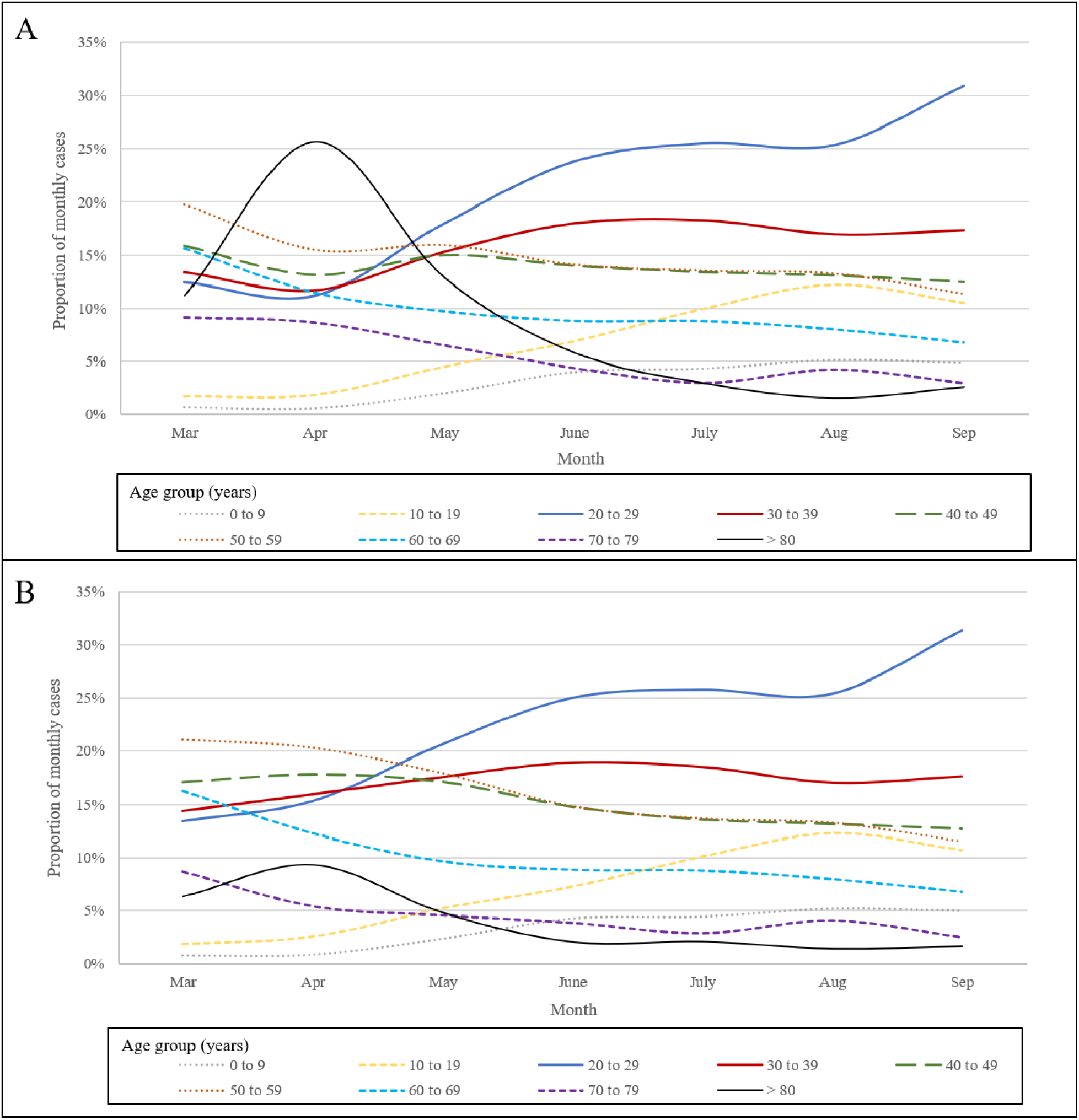
**Distribution of COVID-19 cases by age group from March 1 to September 30, 2020. Figure 1A) (top) shows the distribution of cases by age groups including LTC residents. Figure 1B) (bottom) shows the distribution of cases when LTC residents were excluded from analysis**.

This trend is still evident in the analysis excluding LTC residents (Figure 1B). Starting in June, the age groups 20-39 years account for the greatest proportion of reported cases, with the proportion of cases in the 20-29 year age group increasing from 11% in April to 31% in September.

Analysis of co-morbidities showed a decreasing proportion of cases with the three co-morbidities over time. In March, 11% of all cases were diabetic vs. 4% in September, while cases who were immunocompromised or had renal conditions each decreased from 3% in March to 1% in September. The proportion of cases with neither of these co-morbidities increased from 85% in March to 95% in September.

### Acute care resource use

Hospitalization, ICU admission, and IMV use by age and sex are summarized in Figure 2 (all data in Appendix 7). Overall hospital admission was 10%, with males having a slightly higher proportion of hospitalization (11%) compared to females (9%). The proportion of all reported cases of COVID-19 requiring hospitalization decreased over time: it was highest in March at 21% and decreased to 3% by the end of September. This trend is apparent for all age groups, among the elderly: 70 to 79 years (47% dropping to 18%), 80 years or older (38% dropping to 28%), and also in younger age groups: 40 to 49 years (13% dropping to 2%). On average, individuals were hospitalized 7.8 days after their episode date (7.9 from symptom onset). Hospitalizations for all individuals, and excluding LTC residents, are summarized in Table 2.

**Table 2.** Hospitalizations by age and month between March 1 and September 30, 2020, with and without LTC residents.

|  | **Frequency (proportions)\*** |  |  |  |  |  |  |
| --- | --- | --- | --- | --- | --- | --- | --- |
|  | **All individuals** |  |  |  |  |  |  |
| **Age range (years)** | **March** | **April** | **May** | **June** | **July** | **August** | **September** |
| 0 to 9 | 35 (7%) | 65 (3%) | 37 (2%) | 22 (1%) | 17 (1%) | 20 (2%) | 29 (1%) |
| 10 to 19 |  |  |  |  |  |  |  |
| 20 to 29 |  |  |  |  |  |  |  |
| 30 to 39 | 66 (8%) | 71 (4%) | 43 (3%) | 20 (2%) | 14 (2%) | 17 (3%) | 28 (1%) |
| 40 to 49 | 124 (13%) | 121 (6%) | 80 (5%) | 28 (4%) | 16 (3%) | 18 (4%) | 34 (2%) |
| 50 to 59 | 227 (19%) | 293 (13%) | 143 (9%) | 55 (7%) | 40 (8%) | 19 (4%) | 48 (3%) |
| 60 to 69 | 262 (28%) | 348 (21%) | 179 (18%) | 57 (12%) | 28 (8%) | 30 (11%) | 93 (10%) |
| 70 to 79 | 259 (47%) | 390 (31%) | 207 (31%) | 49 (20%) | 29 (25%) | 25 (18%) | 71 (18%) |
| ≥ 80 | 253 (38%) | 781 (21%) | 357 (28%) | 66 (21%) | 51 (44%) | 19 (36%) | 99 (28%) |
| **Total** | **1,226 (21%)** | **2,069 (14%)** | **1,046 (1%)** | **297 (5%)** | **195 (5%)** | **148 (4%)** | **402 (3%)** |
|  | **All individuals, excluding LTC residents** |  |  |  |  |  |  |
| 0 to 9 | 35 (7%) | 65 (3%) | 36 (2%) | 22 (1%) | 17 (1%) | 20 (2%) | 29 (1%) |
| 10 to 19 |  |  |  |  |  |  |  |
| 20 to 29 |  |  |  |  |  |  |  |
| 30 to 39 | 66 (8%) | 67 (4%) | 42 (3%) | 20 (2%) | 14 (2%) | 17 (3%) | 28 (1%) |
| 40 to 49 | 124 (13%) | 115 (6%) | 79 (5%) | 28 (4%) | 16 (3%) | 18 (4%) | 34 (2%) |
| 50 to 59 | 225 (19%) | 268 (13%) | 136 (9%) | 54 (7%) | 40 (8%) | 19 (4%) | 48 (3%) |
| 60 to 69 | 253 (28%) | 271 (21%) | 155 (18%) | 52 (11%) | 27 (8%) | 28 (11%) | 87 (10%) |
| 70 to 79 | 252 (52%) | 267 (47%) | 164 (41%) | 42 (21%) | 27 (24%) | 24 (18%) | 59 (18%) |
| ≥ 80 | 221 (63%) | 437 (45%) | 227 (54%) | 44 (42%) | 45 (57%) | 17 (37%) | 76 (35%) |
| **Total** | **1,176 (21%)** | **1,490 (14%)** | **839 (10%)** | **262 (5%)** | **186 (5%)** | **143 (4%)** | **361 (3%)** |

\* Proportion of hospitalization is the probability of being hospitalized in that month and is calculated by taking the number of individuals hospitalized over the number of cases in that month.
Some age group frequency and proportions are aggregated to avoid small cells.
**NR**, not reported due to small cell

**Figure 2.**
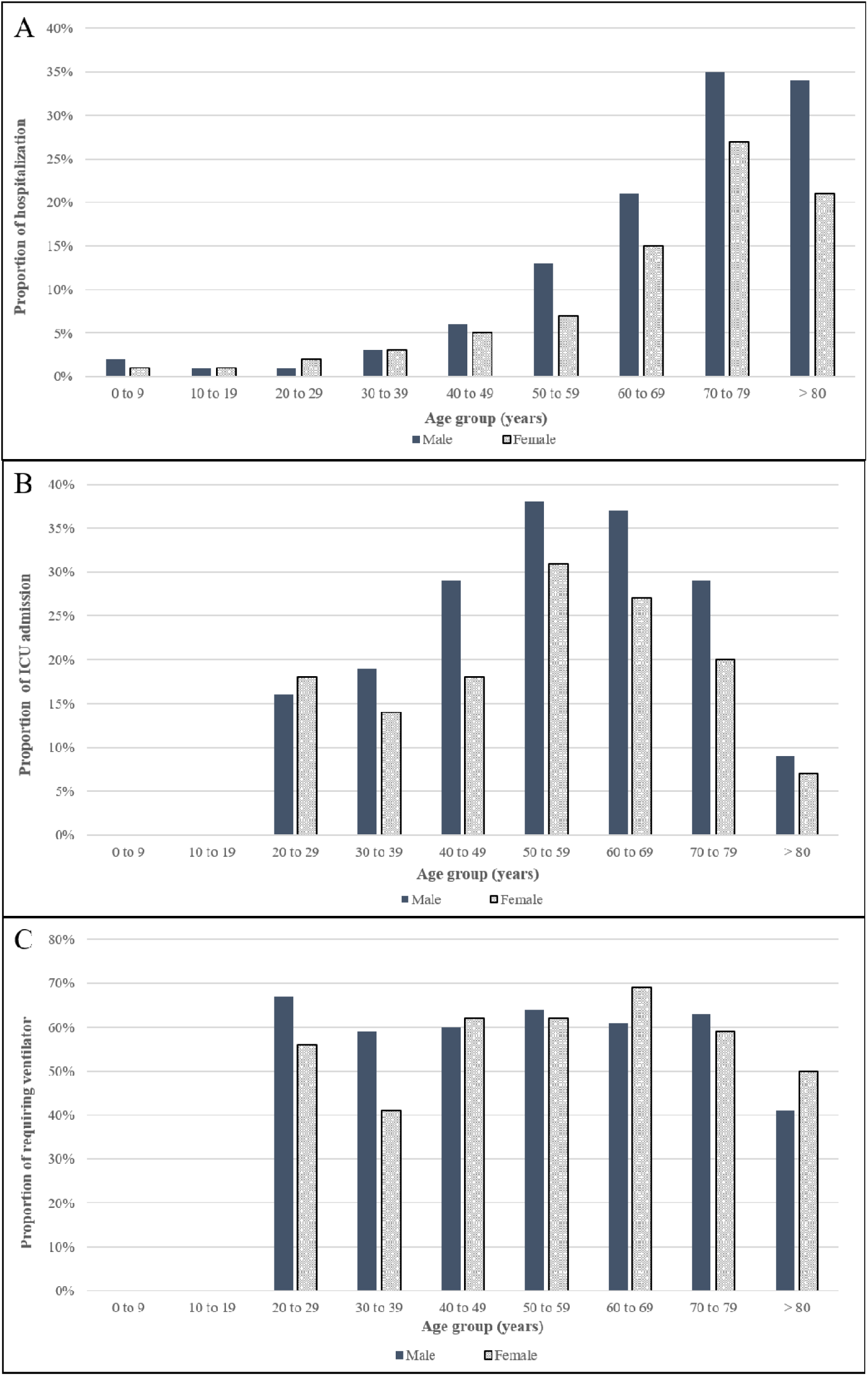
**Acute care resource utilization by age and sex for A) hospitalization B) ICU admissions given hospitalization C) IMV given ICU admission\*** *”Others” group for all, and age 0 to 9, 10 to 19 for Fig 2B, Fig 2C are not shown due to small cells

Analysis of the three comorbidities (Appendix 8) showed that proportion of hospitalization decreased from March to September for cases with diabetes (42% to 14%), immunocompromised (33% to 13%), cases with renal conditions (54% to 36%) and individuals with two or more of these conditions (61% to 42%). However, proportion of ICU admissions were similar or increased in the same period.

Overall, 22% of hospitalized patients required admission to the ICU, with males being more likely to require ICU care (26%) compared to females (17%). ICU admission was highest for males and females between the ages of 50 and 69 years, at 37% and 28% respectively. IMV was required for 60% of COVID-19 cases admitted to ICU. ICU admissions and IMV were highest in March at 32% and 68%, respectively, and decreased over the course of the pandemic to 21%, and 42%, respectively in September. ICU admissions and IMV, with and without LTC residents, are summarized in Appendix 7.

### Length of stay

The mean LOS for those admitted to the ward was 12.8 days. Individuals who required ICU care had an average LOS of 14.6 days (8.5 days in the ICU, and 6.1 days in the ward pre- or post-ICU). Individuals requiring IMV had an average LOS of 29.7 days (20.5 ICU with IMV, 1.2 days in the ICU pre- or post-ventilation, and 7.94 days in the ward pre- or post-ICU care).

The mean LOS by month for each of the three levels of acute care overall, and with LTC residents excluded is presented in Table 3. The number of days spent in the ward for individuals not requiring ICU admission decreased from an average of 16.2 in May to 7.7 days in September. Similarly, the LOS for ICU patients who required IMV decreased from an average of 21.5 days in April to 14.4 days in September.

**Table 3.** Length of stay by level of care between March 1 and September 30, 2020.

|  | **Months, mean (SD) days** |  |  |  |  |  |  |  |
| --- | --- | --- | --- | --- | --- | --- | --- | --- |
|  | **Includes all individuals with outcome of ‘recovered’ or ‘fatal’** |  |  |  |  |  |  |  |
| **Level of Care** | **March** | **April** | **May** | **June** | **July** | **August** | **September** | **7-month average** |
| Ward patients | 10.3 (14.1) | 14.2 (16) | 16.1 (18) | 9.2 (8.5) | 10.4 (12.9) | 6.6 (4.5) | 7.7 (6.1) | 12.8 (15.4) |
| ICU patients, no ventilation | 9.7 (8.7) | 8.3 (8.2) | 7.1 (5.6) | 8.7 (9.1) | 8.5 (4.4) | 12.3 (12.4) | 7.1 (6.7) | 8.5 (7.8) |
| ICU patients, ventilation | 21.5 (18.7) | 20.4 (18.7) | 21 (17.5) | 18 (16.3) | 16 (11.1) | 10.7 (7.3) | 14.4 (8) | 20.5 (18.1) |
|  | **Excludes LTC residents with outcome of ‘recovered’ or ‘fatal’** |  |  |  |  |  |  |  |
| **Level of Care** | **March** | **April** | **May** | **June** | **July** | **August** | **September** | **7-month average** |
| Ward patients | 9.4 (12.6) | 11.7 (14.1) | 14.2 (17.1) | 8.4 (7.7) | 10.3 (13.2) | 6.4 (4.3) | 7.1 (5.7) | 11 (13.8) |
| ICU patients, no ventilator | 9.7 (8.8) | 7.9 (7.6) | 7.2 (5.6) | 9.4 (9.5) | 8.2 (4.5) | 12.3 (12.4) | 7.5 (6.8) | 8.5 (7.7) |
| ICU patients, ventilator | 21.4 (18.7) | 20.3 (17.9) | 21.3 (17.6) | 18.6 (16.5) | 16 (11.1) | 10.7 (7.3) | 14.4 (8) | 20.5 (17.8) |

**ICU**, intensive care unit; **SD**, standard deviation

The LOS distribution for all three levels of acute care are shown in Appendices 9 to 11.

### Mortality

Among cases designated as resolved, the overall mortality during our observation period was 6.2% (3,037 deaths out of 49,362 resolved cases), and 2.4% (1,065 deaths out of 43,500 resolved cases) when excluding LTC residents. Overall mortality is summarized by month in Appendix 12. Mortality decreased from a high of 13% in April to 1% in September for all individuals, and from 5% to 1% when excluding LTC residents.

Age and sex-specific mortality by highest level of acute care are summarized in Table 4. Mortality for individuals not requiring any level of hospitalization, and for individuals requiring acute care was 4% (1,715 deaths out of 44,475 resolved cases), and 27% (1,322 deaths out of 4,887 resolved hospitalizations), respectively. Excluding LTC residents, these proportions were 0.5% (184 deaths out of 39,488 resolved cases) and 22% (881 deaths out of 4,012 resolved hospitalizations), respectively.

**Table 4.**
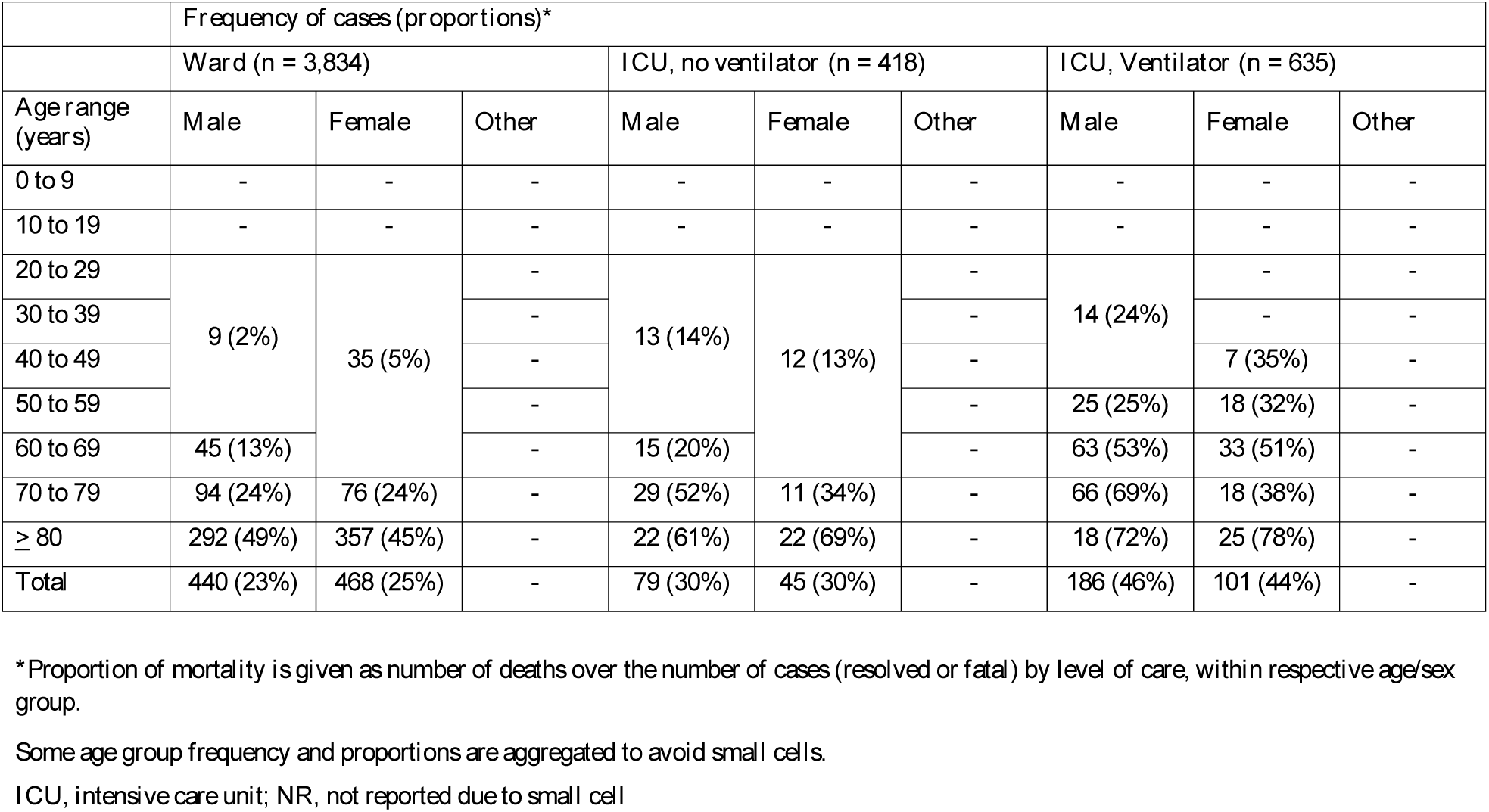
Mortality by level of care, age and sex.

Overall mortality (excluding LTC) for individuals receiving care only in the ward, individuals receiving care in the ICU, and individuals requiring IMV were 24% (17%), 30% (27%), and 45% (44%), respectively. Among COVID-19 patients admitted to the ward, mortality was highest among individuals age 70-79 years (24%) and among those age 80 years and older (47%). Mortality for individuals requiring ICU admission was highest in individuals age 70 to 79 years (43%), and over 80 years (65%). Among patients with COVID-19 who required IMV, mortality was similarly highest amongst the older populations: 54% among those age 70-79 years, and 75% among those over 80 years of age.

### Interpretation

We provide a descriptive analysis of COVID-19 cases from March 1, to September 30, 2020, by age, sex, and LTC residency. The population infected with COVID-19 changed over time: predominantly older age groups in the first three months, and as the summer progressed, infections were predominantly among younger age groups. As the demographics of those infected changed, so too did healthcare resource use – older age groups had a larger proportion of cases requiring admission to hospital, ICU, and need for IMV. The burden of infection among LTC residents resulted in significant morbidity and mortality; LTC residents represented approximately 12% of total cases, and the majority of deaths (65%).

Our results show a decline in the proportion of cases hospitalized, requiring ICU resources, and LOS between March and September. However, the decrease in acute care resource use and mortality was observed overall and within age strata, suggesting that these decreases cannot be entirely explained by changing age distribution over time. There are several potential explanations for these observations. First, changes in clinical practice pattern may have resulted in reduced hospitalizations, utilization of critical care resources and shorter hospital stays. As clinicians gained more experience caring for patients with COVID-19, they may have become comfortable with expectant management, favouring non-invasive oxygenation and/or ventilation inside and outside the ICU, in lieu of early IMV, which may lower prolonged hospitalization and decrease mortality from ventilator associated bacterial pneumonia.(10–13) Further,, data to support the use of prone positioning to improve oxygenation in non-intubated patients, and evidence supporting the use of dexamethasone began to emerge as the pandemic progressed.(14–16) Second, it is possible that mask use or other public health measures including physical distancing, have resulted in lower viral loads among infected individuals, which may be associated with reduced severity of illness.(17) Finally, the finding that there was a decrease in infected individuals who had high-risk co-morbidities over time could be a function of greater awareness and protections in place to protect those at higher risk of severe outcomes, thereby decrease the risk level among the infected cases over time and thus the need for hospitalization and ICU care.

While results from this descriptive study suggest that the proportion of acute care resource use, outcomes (mortality), and LOS are decreasing, it does not imply that the disease has become less severe and does not capture long-term sequelae. A growing body of evidence describes long-term sequelae experienced by many who had mild acute illness, including memory loss and fatigue months after their initial illness, a condition that is being described as “long-COVID”.(18,19)

Our analysis has several limitations. The CCM Plus is an administrative dataset and is subject to underreporting and potential misclassification.(20,21). Testing policies and case definitions have changed between March and September 2020 in Ontario. For example, switch to appointment-only testing in September with stricter criteria means the data may capture more severe cases (and less severe cases), further complicating interpretation of the shifting outcomes. Furthermore, since this data has been collated from various sources, it may be more prone to underreporting of outcomes and data entry errors. Backfilling of data can result in data being added later, delaying the reporting of outcomes up to two months. While we present September outcomes in our analysis, they should be interpreted with caution as hospitalizations and outcomes are lagging indicators (22) i.e., the data is right-censored. This would underestimate all outcomes presented, especially LOS and mortality.

Some of these trends may be confounded by changing healthcare seeking behaviours, public health interventions, implementing and lifting of restrictions, and individual differences (socioeconomic status, co-morbidities, geography)y.(21) As such, we presented data descriptively and believe it would be inappropriate to present statistical measures to identify associations without fully adjusting for confounding.

Despite these limitations, to our knowledge, this study is the first to describe COVID-19 case demographics, acute care use, mortality and LOS stratified by age and sex over seven months of the pandemic in Ontario, Canada. We were able to show the demographics and outcomes over time, capturing the different stages of the pandemic. These insights are critical for policy-makers and capacity planners as the pandemic evolves further.

Centralized public health database like CCM Plus can provide timely population data and can be used in the future, especially if linked to other health administrative data (i.e., hospital discharge data) to understand long-term outcomes of COVID-19 infection. Further, our findings can be used to inform modeling and other studies estimating the impact of COVID-19 and predicting healthcare resource needs.

## Data Availability

The portion derived from government registries are available by requests directed to those agencies.

