## Appendix for "COVID-19 Demographics, Acute Care Resource Use and Mortality by Age and Sex in Ontario, Canada: Population-based Retrospective Cohort Analysis"

### Appendices

#### Appendix 1. CCM Plus variables used for analysis

| Variable Name | Format |
|---|---|
| ACCURATE_EPISODE_DATE | YYYY-MM-DD |
| AGE_GRP | age0_9, age10_19, age20_29, age30_39, age40_49, age50_59, age60_69, age70_79, age80PLUS |
| CASE_CREATED_DATE | YYYY-MM-DD |
| CASE_STATUS_DESCRIPTION | Closed, Open |
| CASE_REPORTED_DATE | YYYY-MM-DD |
| CLIENT_DEATH_DATE | YYYY-MM-DD |
| CLIENT_GENDER2 | Male, Female, Other |
| DIABETES | Yes, No, Unk, FRE |
| DIAGNOSING_HEALTH_UNIT | Public health units (4-digit code) |
| DISPOSITION_DESCRIPTION | Complete, Pending, etc. |
| HOSPADMITDATE | YYYY-MM-DD |
| HOSPDISCHARGEDATE | YYYY-MM-DD |
| HOSPITALIZED | Yes, No |
| ICU | Yes, No |
| ICUENDDATE | YYYY-MM-DD |
| ICUSTARTDATE | YYYY-MM-DD |
| IMMUNOCOMPROMISED | Yes, No, Unk |
| INTUBATION | Yes, No |
| INTUBENDDATE | YYYY-MM-DD |
| INTUBSTARTDATE | YYYY-MM-DD |
| LTCH_RESIDENT | Yes, No |
| OUTCOME1 | Resolved, Fatal, etc. |
| RENALCONDITIONS | Yes, No, Unk |
| SOURCE | CCM, iPHIS, LCMS |
| SPECIMENDATE | YYYY-MM-DD |
| SYMPTOMONSETDATE | YYYY-MM-DD |
| VENTENDDATE | YYYY-MM-DD |
| VENTILATOR | Yes, No |
| VENTSTARTDATE | YYYY-MM-DD |

**Appendix 2. Days between "Case Reported" Date and "Accurate Episode" Date in CCM Plus**

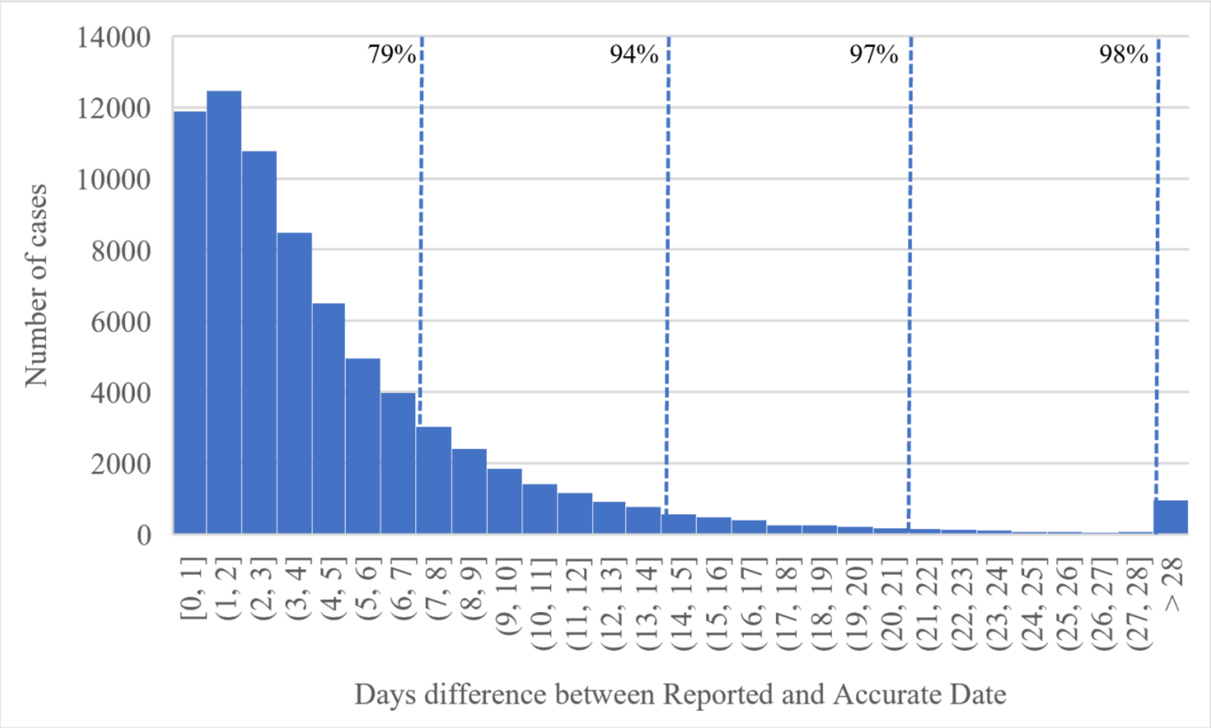

This graph describes the difference in the number of days between the "case reported" date and "accurate episode" date in the CCM plus dataset for all individuals. The dotted lines with percentages represent the cumulative number of cases with up to that difference. From this graph, 94% of cases have an accurate episode date 14 days after their case reported date.

**Appendix 3. Documentation for data clean up**

The following steps were followed to ensure those who used acute care resources are captured. For example, someone who required a ventilator/intubation is counted as 1, and, by definition, required ICU admission. Those who needed ICU admission were, by definition, hospitalized. We assumed that those who are intubated, are counted as individuals who required ventilator use, and to calculate the length of stay for individuals requiring ventilation.

The variables in **bold** were created, whereas other CAPITALIZED variables are from the original data source.

1. Create new variable **VENT**=1 if INTUBATION or VENTILATOR = 'Yes'. This is needed to count those who required ventilation.
2. Create new variable **ICU2** = 1, if ICU = Yes or VENT= 1. This is needed for the ICU proportions data. Some who are ventilated or intubated, have nothing in the ICU data field ('Yes' or 'No') or their start/end dates.
3. Create new variable **HOSP** = 1, if ICU2=1 or HOSPITALIZED= "Yes". This is needed for the hospitalization proportions. Some who are on ventilator, or admitted to ICU have nothing in the HOSPITALIZATION data field or their start/end dates.
4. Create new variable **VENTSTART**: take earliest date from VENTSTARTDATE, INTUBSTARTDATE, VENTENDDATE, INTUBENDDATE. If there are no dates then show a blank.
5. Create new variable **VENTEND**: take latest date from VENTSTARTDATE, INTUBSTARTDATE, VENTENDDATE, INTUBENDDATE. If there are no dates then show a blank.
6. Create new variable **VENTCHECK**: This variable will show 1 if VENT = 1, VENTSTART – VENTEND = 0, and (count(*DATE) =1 or INTUBSTARTDATE=VENTSTARTDATE, else 0. The purpose of this variable is to flag if there is only one date in the four *DATE fields for intubation and ventilation, or if there are two dates but they are equal (e.g., INTUBSTARTDATE = VENTSTARTDATE) and this date is being used for the VENTSTART and VENTEND.
7. Create new variable **VENTEND2**: if ICU2=1, VENTCHECK=1, and count(ICUSTARTDATE, ICUENDDATE) > 1, take ICUENDDATE, otherwise VENTEND. The purpose of this variable is to check if there is a VENTCHECK flag, then see if there is an ICU end date from original dataset. If yes, then assume that for VENTEND2, if not then take the current VENTEND.
8. Create new variable **VENTSTART2**: if ICU2=1, VENTCHECK=1, and count(ICUSTARTDATE, ICUENDDATE) > 1, take ICUSTARTDATE, otherwise VENTSTART. The purpose of this variable is to check if there is a VENTCHECK flag, then see if there is an ICU start date from original dataset. If yes, then assume that for VENTSTART2, if not then take the current VENTSTART.
9. Create a new variable **VENT_LOS**: If VENTEND2-VENTSTART2 < = 0 AND VENTCHECK = 1, then "X", else VENTEND2-VENTSTART2. This variable is used for the LOS for patients on the ventilator. In the first part of that logic statement, it says if there is a flag, and the dates are not in the correct order, then LOS is not calculated and these individuals are excluded.

**Appendix 4. Documentation for data clean up for ICU**

The following steps were used to ensure those who received ventilator use are counted for in the ICU use, and to calculate the length of stay for individuals requiring ICU care.

The variables in **bold** were created, whereas other CAPITALIZED variables are from the original data source.

1. Create new variable **ICUSTART**: if ICU2 = 1, take earliest date from: ICUSTART, ICUEND, VENTSTART2. If there are no dates then show a blank.
2. Create new variable **ICUEND**: if ICU2 = 1, take latest date from: ICUSTART, ICUEND, VENTEND. If there are no dates then show a blank.
3. Create new variable **ICUCHECK**: This variable will show 1 if ICU2 = 1, ICUSTART – ICUEND = 0, and count (ICUSTARTDATE, ICUENDDATE) =1, else 0. Same rationale as VENTCHECK in A2.
4. Create a new variable **ICUEND2:** if HOSP=1, ICUCHECK=1, and count(HOSPADMITDATE, HOSPDISCHARGEDATE) > 1, take max of HOSPADMITDATE/HOSPDISCHARGEDATE, otherwise keep ICUEND. The purpose of this variable is to check if there is a ICUCHECK flag, then see if there is an HOSP end date from original dataset. If yes, then assume that for ICUEND2, if not then take the current ICUEND.
5. Create a new variable **ICUSTART2**: if HOSP=1, ICUCHECK=1, AND COUNT(HOSPADMITDATE, HOSPDISCHARGEDATE) > 1, take min of HOSP*DATE, otherwise ICUSTART. The purpose of this variable is to check if there is a ICUCHECK flag, then see if there is a HOSP start date from original dataset. If yes, then assume that for ICUSTART2, if not then take the current ICUSTART.
6. Create a new variable **ICU_LOS**: If ICUEND2-ICUSTART2 < = 0 AND ICUCHECK = 1, then "X", else ICUEND2-ICUSTART2. IFERROR then "X". This variable is used for the LOS for patients in the ICU. In the first part of that logic statement, it says if there is a flag, and the dates are not in the correct order, then LOS is not calculated and these individuals are excluded.

**Appendix 5. Documentation for data clean up for hospitalizations**

The following steps were used to ensure those who received ventilator use or ICU care are counted for in the hospitalizations, and to calculate the length of stay for individuals requiring hospitalization.

The variables in **bold** were created, whereas other CAPITALIZED variables are from the original data source.

1. Create new variable **HOSPSTART**: if HOSP = 1, take earliest date from: HOSPADMITDATE, HOSPDISCHARGEDATE, ICUSTART2, else blank. If there are no dates then show a blank.

   *The biggest problem with this data field was the HOSPADMITDATEs were frequently in the future of the HOSPDISCHARGEDATE so we needed to make sure chronologically they made sense (the majority).*

2. Create new variable **HOSPEND**: if HOSP = 1, take latest date from: ICUEND2, HOSPADMITDATE, HOSPDISCHARGEDATE, else blank. If there are no dates then show a blank.

3. Create new variable **HOSPCHECK**: if HOSP = 1, HOSPSTART – HOSPEND = 0, and count(HOSPADMITDATE + HOSPDISCHARGEDATE) = 1, then 1, else 0. Same rationale as the other VENTCHECK and ICUCHECK.

4. No need to create **HOSPEND2** because there is nothing left to adjust to at this stage. For those with no discharge date, there is no assumption that can be made here.

   *For all calculations, we only used infected individuals who are considered 'Resolved' or 'Fatal' in the OUTCOME1 field so the analysis would not be affected by censoring.*

5. Create new variable **HOSPSTART2** for those who are missing HOSPSTART (and subsequently HOSPADMITDATE). If HOSPCHECK =1, COUNT(HOSPADMITDATE) = 0, COUNT(SYMPTOMONSETDATE)=1, AND SYMPTOMONSETDATE < HOSPDISCHARGEDATE, then = SYMPTOMONSETDATE, else HOSPSTART. This assumes that for those with a hospitalization, and a discharge date, that their admit date is time of symptom onset if they had no admission date *(assumption).*

6. Create new variable **WARD_LOS**. If HOSPEND-HOSPSTART2 < = 0 and HOSPCHECK = 1, then "X", else if HOSPSTART2 < ACCURATE_EPISODE_DATE, "X", then HOSPEND-HOSPSTART2. The else if statement of code is to correct for HOSPADMIT that were erroneously coded, or before the ACCURATE_EPISODE_DATE.

   *If the person had no date fields completed but were considered hospitalized, they were counted in the proportions but excluded for LOS analysis.*

7. Created two variables **DELAY_Symptom:** if HOSP = 1, Count(SYMPTOMONSETDATE)=1, and HOSPSTART2 >= SYMPTOM_ONSET_DATE, then HOSPSTART2 – SYMPTOM_ONSET_DATE, else "NA". **DELAY_Accurate:** The same was done but using ACCURATE_EPISODE_DATE instead of SYMPTOM_ONSET_DATE.

   *The else if statement was to remove those with dates that were incorrectly entered into the HOSPADMITDATE field of 2018, 2019, early 2020 (could be errors) .*

**Appendix 6. RECORD Checklist**

| | Item No. | STROBE items | Location in manuscript where items are reported | RECORD items | Location in manuscript where items are reported |
|---|---|---|---|---|---|
| **Title and abstract** | | | | | |
| | 1 | (a) Indicate the study's design with a commonly used term in the title or the abstract (b) Provide in the abstract an informative and balanced summary of what was done and what was found | | RECORD 1.1: The type of data used should be specified in the title or abstract. When possible, the name of the databases used should be included.<br><br>RECORD 1.2: If applicable, the geographic region and timeframe within which the study took place should be reported in the title or abstract.<br><br>RECORD 1.3: If linkage between databases was conducted for the study, this should be clearly stated in the title or abstract. | Title page (pg.0)<br><br>Title page (pg. 0)<br><br>Linkage was not conducted |
| **Introduction** | | | | | |
| Background rationale | 2 | Explain the scientific background and rationale for the investigation being reported | Pg. 2 | | |
| Objectives | 3 | State specific objectives, including any prespecified hypotheses | Pg. 2 | | |
| **Methods** | | | | | |
| Study Design | 4 | Present key elements of study | Pg. 2-3 | | |

| | | | | | |
|---|---|---|---|---|---|
| | | design early in the paper | | | |
| Setting | 5 | Describe the setting, locations, and relevant dates, including periods of recruitment, exposure, follow-up, and data collection | Pg. 2-3 | | |
| Participants | 6 | *(a) Cohort study* - Give the eligibility criteria, and the sources and methods of selection of participants. Describe methods of follow-up<br>*Case-control study* - Give the eligibility criteria, and the sources and methods of case ascertainment and control selection. Give the rationale for the choice of cases and controls<br>*Cross-sectional study* - Give the eligibility criteria, and the sources and methods of selection of participants<br>*(b) Cohort study* - For matched studies, give matching criteria and number of exposed and unexposed<br>*Case-control study* - For matched studies, give matching criteria and the number of controls per case | | RECORD 6.1: The methods of study population selection (such as codes or algorithms used to identify subjects) should be listed in detail. If this is not possible, an explanation should be provided.<br>RECORD 6.2: Any validation studies of the codes or algorithms used to select the population should be referenced. If validation was conducted for this study and not published elsewhere, detailed methods and results should be provided.<br>RECORD 6.3: If the study involved linkage of databases, consider use of a flow diagram or other graphical display to demonstrate the data linkage process, including the number of individuals with linked data at each stage. | Pg. 2<br>N/Ap<br>Linkage of databases not conducted |
| Variables | 7 | Clearly define all outcomes, exposures, predictors, potential confounders, and effect modifiers. Give diagnostic | | RECORD 7.1: A complete list of codes and algorithms used to classify exposures, outcomes, confounders, and effect modifiers should be provided. If | Pg. 3 and Appendix 1 |

| | | | | |
|---|---|---|---|---|
| | | criteria, if applicable. | | these cannot be reported, an explanation should be provided. |
| Data sources/ measurement | 8 | For each variable of interest, give sources of data and details of methods of assessment (measurement).<br>Describe comparability of assessment methods if there is more than one group | Pg. 3 and Appendix 1-5 | |
| Bias | 9 | Describe any efforts to address potential sources of bias | Pg. 3-4 | |
| Study size | 10 | Explain how the study size was arrived at | Pg. 3-4 | |
| Quantitative variables | 11 | Explain how quantitative variables were handled in the analyses. If applicable, describe which groupings were chosen, and why | Pg 3-4 and Appendix 2-5 | |
| Statistical methods | 12 | (a) Describe all statistical methods, including those used to control for confounding<br>(b) Describe any methods used to examine subgroups and interactions<br>(c) Explain how missing data were addressed<br>(d) *Cohort study* - If applicable, explain how loss to follow-up was addressed<br>*Case-control study* - If applicable, explain how | Pg. 3-4 | |

| | | | | | |
|---|---|---|---|---|---|
| | | matching of cases and controls was addressed<br>*Cross-sectional study* - If applicable, describe analytical methods taking account of sampling strategy<br>(e) Describe any sensitivity analyses | | | |
| Data access and cleaning methods | | .. | | RECORD 12.1: Authors should describe the extent to which the investigators had access to the database population used to create the study population.<br><br>RECORD 12.2: Authors should provide information on the data cleaning methods used in the study. | Pg. 3-4, Appendix 2-5 |
| Linkage | | .. | | RECORD 12.3: State whether the study included person-level, institutional-level, or other data linkage across two or more databases. The methods of linkage and methods of linkage quality evaluation should be provided. | No linkage conducted |
| **Results** | | | | | |
| Participants | 13 | (a) Report the numbers of individuals at each stage of the study (*e.g.*, numbers potentially eligible, examined for eligibility, confirmed eligible, included in the study, completing follow-up, and analysed) | | RECORD 13.1: Describe in detail the selection of the persons included in the study (*i.e.*, study population selection) including filtering based on data quality, data availability and linkage. The selection of included persons can be described in the text and/or by | Pg. 4 |

| | | | | |
|---|---|---|---|---|
| | | (b) Give reasons for non-participation at each stage.<br><br>(c) Consider use of a flow diagram | | means of the study flow diagram. |
| Descriptive data | 14 | (a) Give characteristics of study participants (*e.g.*, demographic, clinical, social) and information on exposures and potential confounders<br><br>(b) Indicate the number of participants with missing data for each variable of interest<br><br>(c) *Cohort study* - summarise follow-up time (*e.g.*, average and total amount) | Pg. 4-5, Table 1, Figure 1 | |
| Outcome data | 15 | *Cohort study* - Report numbers of outcome events or summary measures over time<br><br>*Case-control study* - Report numbers in each exposure category, or summary measures of exposure<br><br>*Cross-sectional study* - Report numbers of outcome events or summary measures | Pg. 5-7, Table 2-4, Figure 2, Appendix 7-13 | |
| Main results | 16 | (a) Give unadjusted estimates and, if applicable, confounder-adjusted estimates and their precision (e.g., 95% confidence interval). Make clear which confounders were adjusted for and why they were included | Pg. 5-7, Table 2-4, Figure 2, Appendix 7-13 | |

| | | | | |
|---|---|---|---|---|
| | | (b) Report category boundaries when continuous variables were categorized<br><br>(c) If relevant, consider translating estimates of relative risk into absolute risk for a meaningful time period | | |
| Other analyses | 17 | Report other analyses done—e.g., analyses of subgroups and interactions, and sensitivity analyses | Pg. 5-7, Appendix 7-13 | |
| **Discussion** | | | | |
| Key results | 18 | Summarise key results with reference to study objectives | Pg. 7-8 | |
| Limitations | 19 | Discuss limitations of the study, taking into account sources of potential bias or imprecision. Discuss both direction and magnitude of any potential bias | Pg. 8-9 | RECORD 19.1: Discuss the implications of using data that were not created or collected to answer the specific research question(s). Include discussion of misclassification bias, unmeasured confounding, missing data, and changing eligibility over time, as they pertain to the study being reported. |
| Interpretation | 20 | Give a cautious overall interpretation of results considering objectives, limitations, multiplicity of analyses, results from similar studies, and other relevant evidence | Pg. 7-9 | |
| Generalisability | 21 | Discuss the generalisability (external validity) of the study | Pg. 9 | |

| | | | | | |
|---|---|---|---|---|---|
| | | results | | | |
| **Other Information** | | | | | |
| Funding | 22 | Give the source of funding and the role of the funders for the present study and, if applicable, for the original study on which the present article is based | Title page (Pg. 0) | | |
| Accessibility of protocol, raw data, and programming code | | .. | | RECORD 22.1: Authors should provide information on how to access any supplemental information such as the study protocol, raw data, or programming code. | Throughout (Appendix #s referenced) |

**Appendix 7. Hospitalization, ICU admission, and ventilation by age and sex between March 1, 2020 to September 30, 2020**

| | **Frequency of cases (proportions)*** | | | | | | | | |
|---|---|---|---|---|---|---|---|---|---|
| | **Hospitalization (n=5,383)** | | | **ICU Admission (n=1,183)** | | | **Ventilation (n=712)** | | |
| **Age range (years)** | **Male** | **Female** | **Other** | **Male** | **Female** | **Other** | **Male** | **Female** | **Other** |
| 0 to 9 | 12 (2%) | 8 (1%) | | | | | | | |
| 10 to 19 | 21 (1%) | 18 (1%) | | 19 (17%) | 17 (15%) | | 10 (53%) | 9 (56%) | |
| 20 to 29 | 77 (1%) | 88 (2%) | | | | | | | |
| 30 to 39 | 140 (3%) | 119 (3%) | | 27 (19%) | 17 (14%) | | 16 (59%) | 7 (41%) | |
| 40 to 49 | 235 (6%) | 185 (5%) | 8 (3%) | 68 (29%) | 34 (18%) | NR | 41 (59%) | 21 (62%) | NR |
| 50 to 59 | 497 (13%) | 327 (7%) | | 188 (38%) | 100 (31%) | | 120 (64%) | 62 (62%) | |
| 60 to 69 | 602 (21%) | 394 (15%) | | 222 (37%) | 105 (27%) | | 136 (61%) | 72 (69%) | |
| 70 to 79 | 589 (35%) | 440 (27%) | | 169 (29%) | 86 (20%) | | 106 (63%) | 51 (59%) | |
| ≥ 80 | 713 (34%) | 910 (21%) | | 63 (9%) | 66 (7%) | | 26 (43%) | 33 (50%) | |
| **Total** | **2,886 (11%)** | **2,489 (9%)** | **8 (3%)** | **756 (26%)** | **425 (17%)** | **NR** | **455 (60%)** | **255 (60%)** | **NR** |

*Proportion of hospitalization is calculated as number of hospitalizations per number of cases within respective group. Proportion of ICU admission is given as number of ICU admissions per number of hospitalized individuals within respective group. Proportion of ventilation is given as number of individuals requiring ventilation over number of individuals admitted to ICU within respective group.

Some age group frequency and proportions are aggregated to avoid small cells.

ICU, intensive care unit; NR, not reported due to small cell

**Appendix 8. Proportion of hospitalizations and ICU admissions for co-morbid cases between March and September 2020**

| | Frequency (proportions)* | | | | | | |
|---|---|---|---|---|---|---|---|
| | **Hospitalization** | | | | | | |
| **Comorbidity** | **March** | **April** | **May** | **June** | **July** | **August** | **September** |
| Diabetes | 269 (42%) | 440 (29%) | 219 (24%) | 73 (19%) | 51 (19%) | 36 (15%) | 77 (14%) |
| Immunocompromised | 56 (33%) | 94 (35%) | 63 (41%) | 11 (18%) | 10 (22%) | 18 (13%) | |
| Renal conditions | 109 (54%) | 190 (43%) | 67 (36%) | 28 (35%) | 14 (29%) | 9 (24%) | 35 (36%) |
| 2 or more | 68 (61%) | 115 (49%) | 51 (52%) | 19 (50%) | 10 (56%) | 27 (42%) | |
| | **Intensive care unit (ICU) admission** | | | | | | |
| **Comorbidity** | **March** | **April** | **May** | **June** | **July** | **August** | **September** |
| Diabetes | 102 (38%) | 100 (23%) | 57 (26%) | 17 (23%) | 17 (33%) | 9 (25%) | 22 (29%) |
| Immunocompromised | 19 (34%) | 20 (21%) | 13 (21%) | 6 (29%) | | 7 (39%) | |
| Renal conditions | 38 (35%) | 35 (18%) | 27 (40%) | 10 (36%) | 6 (23%) | | 13 (37%) |
| 2 or more | 26 (38%) | 25 (22%) | 19 (37%) | 7 (24%) | | 13 (48%) | |

*Proportions are calculated by taking the number of co-morbid cases hospitalized based on the total number of co-morbid cases by month based on episode date for hospitalizations, and by taking the number of co-morbid cases requiring ICU admission based on the total number of co-morbid cases hospitalized.

Some months do not show exact frequency and show <6 or are merged to show an average over two months because of small cells.

**Appendix 9. Acute care resource utilization by level of care and month between March 1 and September 30, 2020**

| Level of care | March | April | May | June | July | August | September |
|---|---|---|---|---|---|---|---|
| Including all individuals, frequency (proportions)* | | | | | | | |
| Hospitalization, n=5,383 | 1,226 (21%) | 2,069 (14%) | 1046 (10%) | 297 (5%) | 195 (5%) | 148 (4%) | 402 (3%) |
| ICU, no ventilation, n=1,183 | 398 (32%) | 362 (17%) | 197 (19%) | 64 (22%) | 50 (26%) | 29 (20%) | 83 (21%) |
| ICU, ventilation, n=712 | 269 (68%) | 239 (66%) | 106 (54%) | 31 (48%) | 17 (34%) | 15 (52%) | 35 (42%) |
| Excluding those from LTC outbreaks, frequency (proportions)* | | | | | | | |
| Hospitalization, n=4,457 | 1,176 (21%) | 1,490 (14%) | 839 (10%) | 262 (5%) | 186 (5%) | 143 (4%) | 361 (3%) |
| ICU, no ventilation, n=1,113 | 394 (34%) | 318 (21%) | 186 (22%) | 59 (23%) | 48 (26%) | 29 (20%) | 79 (22%) |
| ICU, ventilation, n=681 | 266 (68%) | 215 (68%) | 104 (56%) | 29 (49%) | 17 (35%) | 15 (52%) | 35 (44%) |

* Proportion of hospitalizations are number of hospitalizations over the total number of cases for that month by “case created” date. Proportion of ICU, no ventilation, is the number of ICU admissions for those not needing ventilators over the number of hospitalizations for that month. Proportion of ICU, ventilation is the number of individuals requiring ventilators over the total number of individuals admitted to the ICU during that month.

ICU, intensive care unit; LTC, long-term care

**Appendix 10. Distribution of length of stay for patients hospitalized in the ward for all individuals between March 1 and September 30, 2020**

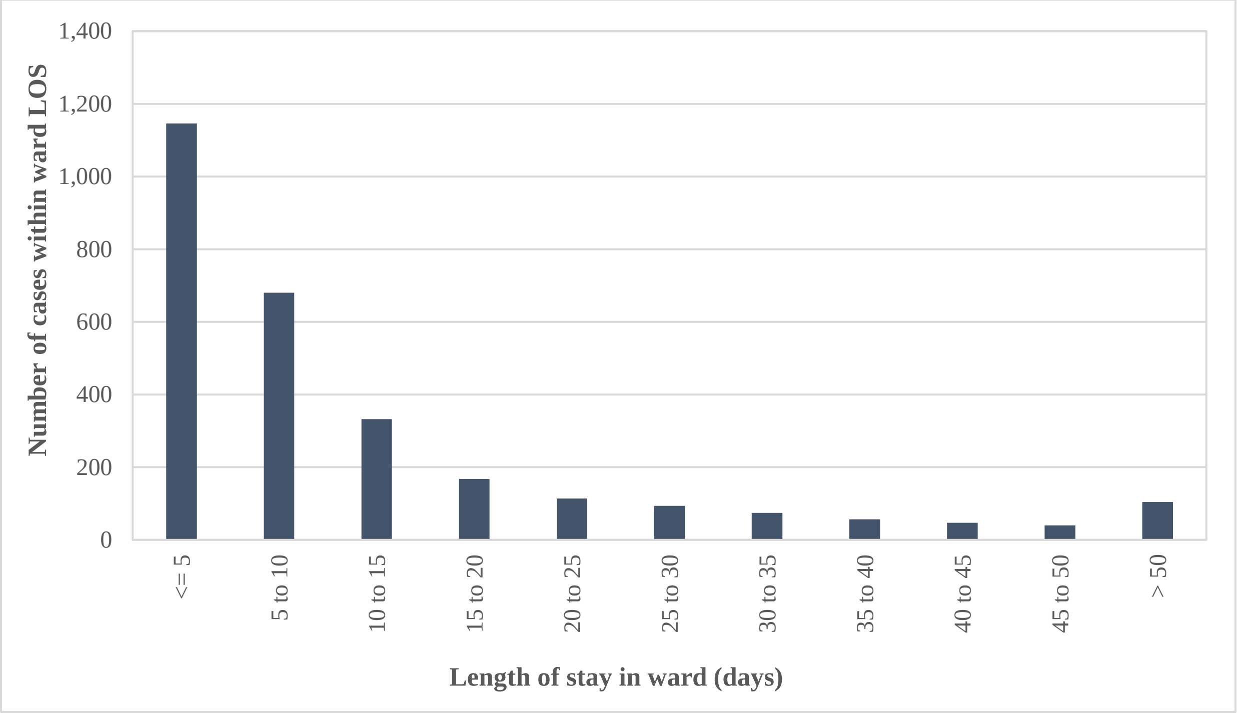

**Appendix 11. Distribution of length of stay for patients admitted to intensive care unit (ICU) care for all individuals between March 1 and September 30, 2020**

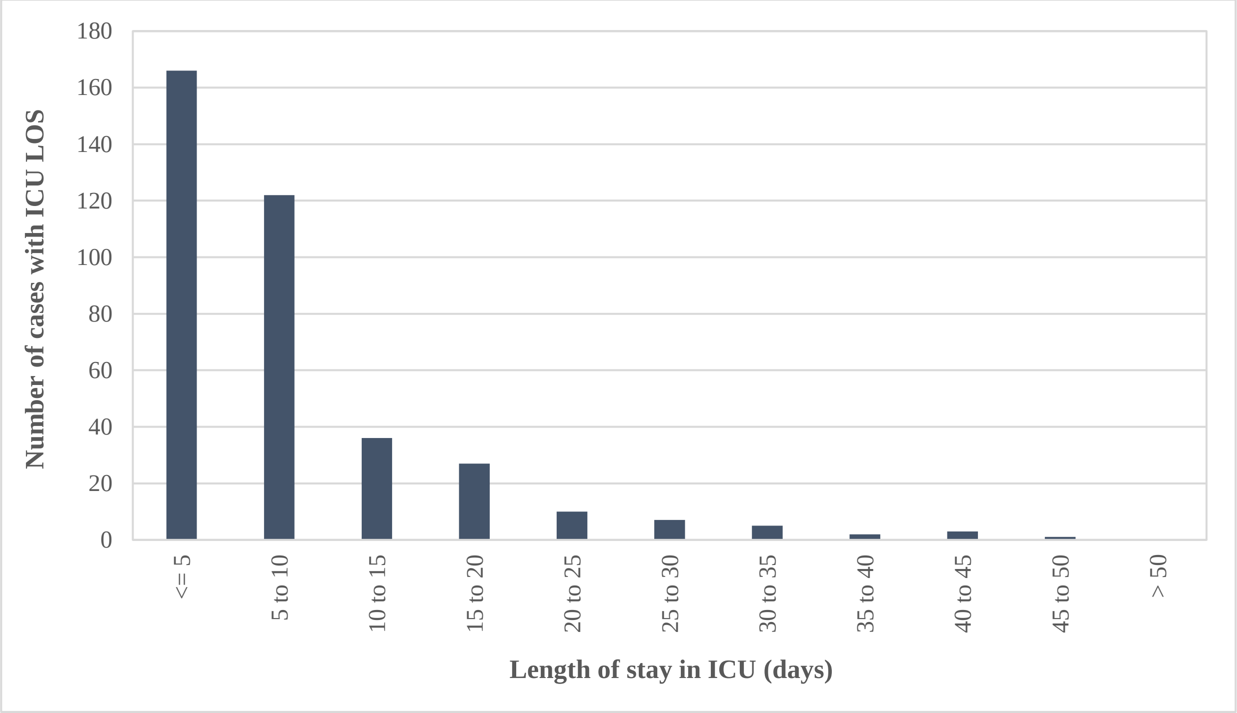

**Appendix 12. Distribution of length of stay for patients requiring ventilation for all individuals between March 1 and September 30, 2020**

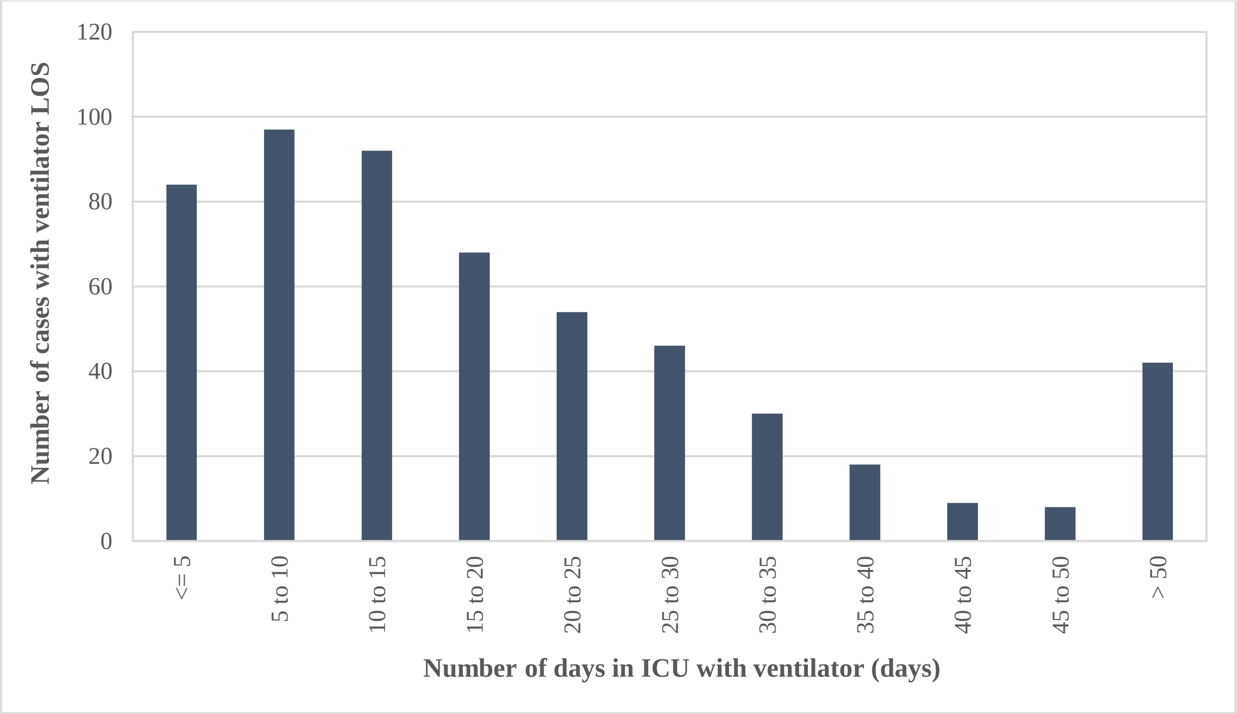

**Appendix 13. Mortality by month between March 1 and September 30, 2020**

| Level of care | March | April | May | June | July | August | September |
|---|---|---|---|---|---|---|---|
| Including all individuals | | | | | | | |
| Frequency (proportion)* | 457 (7.9%) | 1,818 (13.4%) | 514 (5.4%) | 100 (1.9%) | 33 (0.9%) | 17 (0.6%) | 98 (1.1%) |
| Excluding individuals from long-term care residents | | | | | | | |
| Frequency (proportion)* | 271 (5%) | 484 (5%) | 174 (2%) | 43 (1%) | 25 (1%) | 13 (<1%) | 55 (1%) |

* Proportion is calculated by taking deaths as the numerator of all individuals who had an episode date in that month over the total number infected within that month using the episode date
